# Multimodal biomarker characterization of amnestic objective subtle cognitive decline

**DOI:** 10.1101/2025.10.27.25338711

**Authors:** David López-Martos, Raffaele Cacciaglia, Marc Suárez-Calvet, Gemma Salvadó, Mahnaz Shekari, Armand González-Escalante, Marta Milà-Alomà, Anna Brugulat-Serrat, Carolina Minguillon, Matteo Tonietto, Edilio Borroni, Gregory Klein, Clara Quijano-Rubio, Gwendlyn Kollmorgen, Henrik Zetterberg, Kaj Blennow, Juan Domingo Gispert, Oriol Grau-Rivera, Gonzalo Sánchez-Benavides, the ALFA study

**Author notes:** Corresponding authors: David López-Martos, MSc, Barcelonaβeta Brain Research Center (BBRC), Pasqual Maragall Foundation. Wellington 30, 08005, Barcelona, Spain, Oriol Grau-Rivera, MD, PhD, Barcelonaβeta Brain Research Center (BBRC), Pasqual Maragall Foundation. Wellington 30, 08005, Barcelona, Spain, Gonzalo Sánchez-Benavides, PhD., Barcelonaβeta Brain Research Center (BBRC), Pasqual Maragall Foundation. Wellington 30, 08005, Barcelona, Spain. The complete list of collaborators of the ALFA study can be found in the acknowledgments section.

## Abstract

**Background:** Alzheimer’s disease (AD) diagnostic guidelines acknowledge transitional decline prior to clinical onset, yet the absence of practical recommendations for defining this stage presents a major challenge for early detection and risk stratification. Objectively defined subtle cognitive decline (obj-SCD) is increasingly recognized, yet optimal methods for its definition remain uncertain, and consequently, the pathological and neuronal characteristics of this stage remain largely unexplored. The objective of this study was to provide a multimodal biomarker characterization of amnestic obj-SCD, defined using clinically grounded standardized longitudinal neuropsychological criteria in aging and preclinical AD.

**Methods:** This prospective observational study analyzed 3-year longitudinal data from the Alzheimer’s and Families+ (ALFA+) cohort, including cognitively unimpaired participants with available baseline CSF biomarker measurements (normal or AD *continuum* profiles) and longitudinal neuropsychological assessments (2 time points, 3-year follow-up). The primary study measurement, amnestic obj-SCD, was defined using robust longitudinal neuropsychological references combined with multivariate base rate thresholds of significant cognitive decline (Free and Cued Selective Reminding Test, Memory Binding Test, Wechsler Memory Scale IV: Logical Memory). Study outcomes included plasma p-tau217, NfL, and GFAP; CSF p-tau181/Aβ42, NfL, and GFAP; Aβ and tau PET; and Grey Matter volume (GMv) measurements. The associations of amnestic obj-SCD with fluid (plasma and CSF) and neuroimaging (PET and GMv) biomarkers were evaluated using mixed-effects and voxel-wise linear regression models, respectively.

**Results:** A total of 350 CU individuals were included (mean age 61 years; 60% female; mean education, 14 years, 35% CSF Aβ-positive). Applying the criteria defined for clinical staging, amnestic obj-SCD was identified in 10% of the sample. Multimodal biomarker characterization demonstrated significant associations of amnestic obj-SCD with core AD biomarkers, including diagnostic core 1 (higher plasma p-tau217 concentration, higher CSF p-tau181/Aβ42 ratio, and higher global Aβ PET burden) and prognostic core 2 AD biomarkers (higher MTL tau PET burden). Amnestic obj-SCD was also significantly associated with biomarkers of non-specific processes involved in AD pathophysiology, including neurodegeneration (higher plasma and CSF NfL concentrations, reduced GMv in the cingulate cortex, and longitudinal GMv bilateral reductions in the hippocampus) alongside inflammation (higher plasma and CSF GFAP concentrations, and longitudinal GMv increases in neocortical brain regions).

**Discussion:** This study suggests that amnestic obj-SCD reflects early AD-related neuropathological changes (Aβ and tau) and key downstream mechanisms involved in AD pathophysiology (neurodegeneration and inflammation). These findings highlight the need for standardized clinical staging criteria to enhance early detection and risk stratification in aging and preclinical AD.

## Background

Alzheimer’s disease (AD) encompasses a preclinical stage preceding mild cognitive impairment (MCI) and dementia, defined by underlying neuropathological changes without clinical impairment.^1^ In 2011, the National Institute on Aging and the Alzheimer’s Association (NIA-AA) defined subtle cognitive decline as a transitional stage between normal cognition and MCI, characterized by low cognitive performance not meeting clinical thresholds.^2^ The 2018 NIA-AA guidelines^3^ and 2024 Alzheimer’s Association revised criteria^4^ further conceptualized this stage as transitional decline (Stage 2), between the asymptomatic period (Stage 1) and cognitive impairment (Stage 3). Diagnostic guidelines specify that transitional decline should be documented by subtle, subclinical cognitive and/or behavioral decline (1-3 years), based on objective neuropsychological testing and/or subjective reports of decline.

The literature suggests that the most relevant manifestation of Stage 2 may be an objective episodic memory decline, reflecting a subclinical amnestic variant in preclinical AD.^5,6^ In contrast, Subjective cognitive decline (SCD) may not capture this transitional stage, as it is non-specific for AD and may be reduced or absent in MCI patients with impaired cognitive awareness (*i.e.*, anosognosia),^7,8^ limiting its predictive value across the preclinical stage, where distinct metacognitive profiles and trajectories, ranging from heightened to reduced cognitive awareness, have been identified.^9,10^ Although the Alzheimer’s Association^4^ and the International Working Group^11^ provided relevant conceptual frameworks, practical clinical staging guidelines remain lacking.^12,13^ The absence of common standards for defining transitional decline presents a major challenge for early detection and risk stratification.

Clinical research increasingly supports the conceptualization of objectively defined subtle cognitive decline (obj-SCD), reflecting growing interest in this field (see Thomas & Edmonds, 2025, for a systematic review).^14^ These investigations proposed discrete categorizations of obj-SCD consistent with clinical Stage 2, however methodologies varied widely: ranging from single tests to composite scores, distinct cut-offs, cross-sectional versus longitudinal designs, and spanning from clinically based to data-driven approaches. Particularly, longitudinal studies are still few and optimal methods for its definition remain uncertain. While prospective studies show that AD pathophysiology tracks cognitive decline in cognitively unimpaired (CU) individuals,^15,16^ establishing standards for distinguishing normal cognitive trajectories from variability associated with underlying neuropathological progression remains challenging.^17^

Robust neuropsychological references that account for longitudinal clinical progression and/or AD biomarkers provide higher thresholds for defining normal cognition.^18–21^ Established methods for measuring neuropsychological change, such as the Reliable Change Index (RCI) and Standardized Regression-Based (SRB) models, account for test-retest correlation, practice effects, and sociodemographic adjustments shaping intra-individual normative trajectories.^22–24^ Previously, we combined these methods to develop robust AD biomarker-based longitudinal neuropsychological references, demonstrating greater sensitivity in detecting significant cognitive decline, particularly in episodic memory.^25^ The integration of these robust longitudinal references with multivariate base rate thresholds, identifying significant deviations from the normative frequency of low performance across multiple neuropsychological measures,^26,27^ provides a standardized framework for clinical staging defining obj-SCD.

Building on this framework, this study aimed to provide a comprehensive multimodal biomarker characterization of amnestic obj-SCD in aging and preclinical AD, evaluating associations with core AD biomarkers (Aβ and tau) and biomarkers of non-specific processes involved in AD pathophysiology (inflammation and neurodegeneration), ranging from fluid (plasma and cerebrospinal fluid [CSF]) to neuroimaging measurements (positron emission tomography [PET] and Magnetic Resonance Imaging [MRI] Grey Matter Volume [GMv]).

## Methods

### Study Participants

This study was conducted within the Alzheimer’s and Families+ (ALFA+) cohort, a longitudinal extension of the ALFA parent cohort at the Barcelonaβeta Brain Research Center, Barcelona, Spain.^28^ The ALFA cohort recruited CU individuals aged 45–74 years, free from major medical, psychiatric, or neurological conditions. The ALFA+ cohort represents a selected subset of ALFA participants enriched for AD risk factors, including parental history of AD and carriage of the apolipoprotein E (*APOE*) ε4 allele. Eligibility criteria for ALFA+ included prior participation in ALFA and willingness to participate in clinical and neuropsychological assessment, blood and CSF collection, as well as MRI and PET imaging. Participants with cognitive impairment, systemic illnesses, or monogenic AD mutation were excluded. Full details are provided in the Supplementary Material.

In this study, we included CU participants from ALFA+ with available baseline CSF biomarker determination and longitudinal neuropsychological evaluations (two visits over a 3-year follow-up). Individuals with normal CSF biomarker profiles or within the AD *continuum* were retained, whereas individuals with suspected non-AD pathology were excluded, according to the AT(N) classification system (see Multimodal Biomarker Panel and Classification System). Selection criteria were consistent with previous work in the ALFA+ cohort, as previously described. ^25^

### Standard Protocol Approvals, Registrations, and Patient Consents

The ALFA+ study (ALFA-FPM-0311) was approved by the Independent Ethics Committee of Parc de Salut Mar, Barcelona, and is registered on Clinicaltrials.gov (NCT02485730). All participants provided written informed consent, as approved by the Ethics Committee. The study was performed in accordance with the principles of the Declaration of Helsinki.

### Biomarker Measurements

#### Plasma and Cerebrospinal Fluid (CSF)

Participants underwent longitudinal fasting blood and CSF collection.^29,30^ Plasma biomarkers included p-tau217, NfL, and GFAP, while CSF biomarkers included Aβ42/40, p-tau181, t-tau, p-tau181/Aβ42, NfL, and GFAP. All biomarkers except plasma p-tau217 were analyzed at the Clinical Neurochemistry Laboratory, University of Gothenburg, Sweden. Plasma p-tau217 was measured by Eli Lilly using an in-house Meso Scale Discovery (MSD) assay.^31^ CSF Aβ42/40, NfL, and GFAP, as well as plasma NfL and GFAP, were quantified with the NeuroToolKit, a panel of prototype electrochemiluminescence Elecsys® immunoassays on fully automated Cobas® e 411 or e 601 modules (Roche Diagnostics International Ltd, Rotkreuz, Switzerland). CSF p-tau181 and t-tau were measured with Elecsys Phospho-Tau (181P) and Total-Tau electrochemiluminescence immunoassays on Cobas e 601 module, and CSF p-tau181/Aβ42 with the same Elecsys platform.

#### Positron Emission Tomography (PET)

Participants underwent longitudinal Aβ PET, with a subset also completing tau PET at follow-up. Scans were acquired on a Siemens Biograph mCT scanner with cranial CT for attenuation correction. Aβ PET (∼185 MBq [¹⁸F]flutemetamol) was acquired 90–110 min post-injection, and tau PET (368.75 MBq [¹⁸F]RO948) 70–90 min post-injection (20 min, 4 × 5 min frames for both Aβ and tau PET). Images were reconstructed using ordered subset expectation maximization with time-of-flight and point spread function modeling (Aβ: 8 iterations, 21 subsets, 3 mm; tau: 4 iterations, 21 subsets, 4 mm), co-registered to T1-weighted MRI, and spatially normalized to MNI space via DARTEL.^32^ Standardized Uptake Value Ratios (SUVRs) were defined using the whole cerebellum as the reference for Aβ PET and inferior cerebellum for tau PET. Longitudinal Aβ PET changes were computed voxel-wise. All images were smoothed with an 8-mm FWHM Gaussian kernel. Additionally, Aβ PET Centiloid units were quantified using validated methods.^33^

#### Magnetic Resonance Imaging (MRI)

Participants underwent longitudinal MRI scans on a 3T Philips Ingenia CX scanner using T1-weighted 3D-turbo field echo sequences (0.75 mm³ isotropic voxel; FOV 240×240×180 mm³; flip angle 8°; TR 9.9 ms; TE 4.6 ms; TI 900 ms). Grey matter (GM) segmentation was performed using SPM12 and mapped to common space, with a template generated from these segmentations warped to MNI space via DARTEL.^32^ Native GM images were modulated by Jacobian determinants to preserve local GM volume (GMv).^34^ Longitudinal GMv changes were computed using SPM’s pairwise longitudinal registration in native space, and resulting Jacobians were normalized to MNI space using DARTEL. All images were smoothed with an 8-mm FWHM Gaussian kernel. Total intracranial volume (TIV) was computed as the sum of GM, white matter (WM), and CSF volumes.

#### Multimodal Biomarker Panel and Classification System

According to previous work in the ALFA+ cohort, the AT(N) classification system was used to define biomarker profiles in consistency with the 2018 NIA-AA research framework.^3^ CSF biomarker classification was determined using previously validated cut-offs in the ALFA+ cohort: Aβ-positivity (A+) was defined by Aβ42/40 ratio < 0.071, tau-positivity (T+) by p-tau181 > 24 pg/mL, and neurodegeneration-positivity ([N]+) by t-tau > 300 pg/mL. ^29^ Following 2024 Alzheimer’s Association revised criteria,^4^ this study focused on core AD biomarkers and biomarkers of non-specific processes involved in AD pathophysiology, including plasma, CSF, PET, and MRI biomarker measurements. Full details are provided in the Supplementary Material (Supplementary Tables 1 and 2).

**Table 1.** Baseline Characteristics of the Participants.

| Characteristics | Full Sample,<br>n= 350 (100%) | Stable,<br>n= 315 (90%) | Amnestic obj-SCD,<br>n= 35 (10%) | *p*-value |
| --- | --- | --- | --- | --- |
| *Demographic* |  |  |  |  |
| Age, mean (SD) | 60.93 (4.72) | 60.59 (4.62) | 63.94 (4.61) | **< 0.001** |
| Female sex, n (%) | 211 (60.29) | 191 (60.63) | 20 (57.14) | 0.827 |
| Education (years), mean (SD) | 13.53 (3.48) | 13.59 (3.46) | 12.91 (3.66) | 0.274 |
| *Genetic* |  |  |  |  |
| *APOE-ε4* carriers, n (%) | 191 (54.57) | 169 (53.65) | 22 (62.86) | 0.390 |
| *Biomarkers* |  |  |  |  |
| Aβ-positive status, n (%) | 122 (34.86) | 101 (32.06) | 21 (60) | **0.002** |
| Plasma p-tau217 (pg/ml), mean (SD) | 0.153 (0.090) | 0.149 (0.087) | 0.191 (0.107) | **0.009** |
| CSF p-tau181/Aβ42 (hybrid ratio), mean (SD) | 0.014 (0.011) | 0.013 (0.082) | 0.024 (0.022) | **< 0.001** |
| Aβ PET (Centiloids), mean (SD) | 3.52 (17.25) | 1.77 (14.77) | 18.93 (27.44) | **< 0.001** |
| *Neuropsychology* |  |  |  |  |
| MMSE, mean (SD) | 29.18 (0.92) | 29.21 (0.90) | 28.89 (1.05) | **0.048** |
| SCD Participant, positive status, n (%) | 98 (28) | 83 (26.35) | 15 (42.86) | 0.062 |
| SCD-Q Participant, total, mean (SD) | 4.11 (4.38) | 3.88 (4.25) | 6.18 (5.08) | **0.004** |
| SCD Study Partner, positive status, n (%) | 16 (4.57) | 12 (3.81) | 4 (11.43) | 0.064 |
| SCD-Q Study Partner, total, mean (SD) | 1.53 (2.47) | 1.40 (2.33) | 2.71 (3.26) | **0.003** |

Results presented correspond to the mean and standard deviation (SD) for continuous variables and number of observations (n) and percentage (%) for categorical variables. Comparisons between stable and amnestic obj-SCD groups were performed using ANOVA, chi-square ( $\chi^2$ ), or Fisher's Exact Test, depending on the type of variable and distribution of the data, with corresponding nominal $p$ -values reported. Amnestic obj-SCD was defined using robust longitudinal neuropsychological references with a multivariate base rate threshold of significant cognitive decline. Episodic memory was evaluated with standardized neuropsychological tests (Free and Cued Selective Reminding Test; Memory Binding Test; WMS-IV: Logical Memory Subtest). The previously validated biomarker cut-off used to define Aβ-positive status in the ALFA+ cohort was < 0.071 for the CSF Aβ42/40 ratio. Bold text indicates a significant association ( $p$ -values < 0.05). Aβ : β-amyloid; APOE: apolipoprotein E; CSF: cerebrospinal fluid; MMSE: mini-mental state examination; PET: positron emission tomography; p-tau: phosphorylated tau; SCD: subjective cognitive decline; SCD-Q: subjective cognitive decline questionnaire.

**Table 2.**
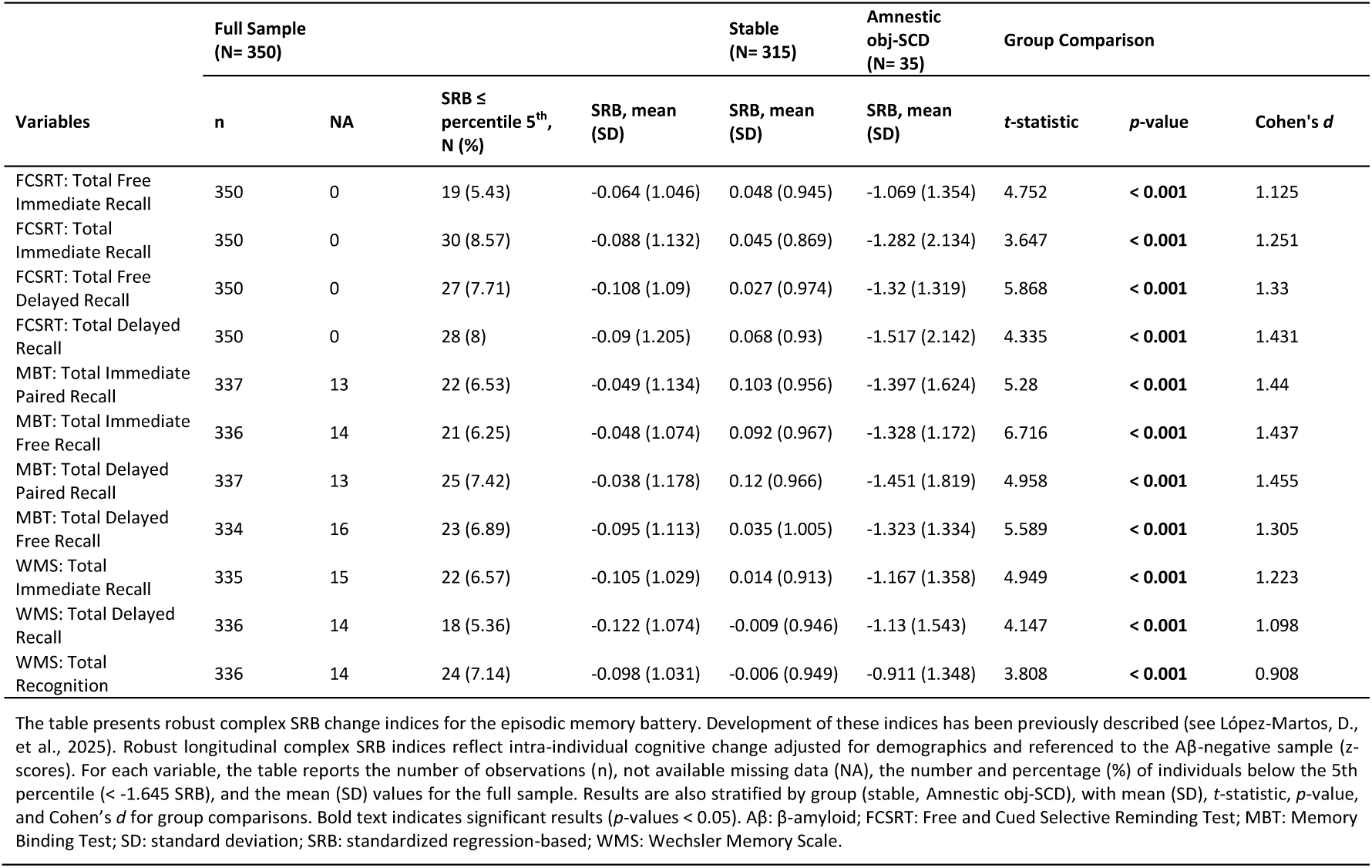
AD Biomarker-based Robust Complex SRB Change Indices for the Episodic Memory Battery.

| Variables | Full Sample<br>(N= 350) |  |  |  | Stable<br>(N= 315) | Amnesic<br>obj-SCD<br>(N= 35) | Group Comparison |  |  |
| --- | --- | --- | --- | --- | --- | --- | --- | --- | --- |
| | n | NA | SRB ≤<br>percentile 5<sup>th</sup>,<br>N (%) | SRB, mean<br>(SD) | SRB, mean<br>(SD) | SRB, mean<br>(SD) | t-statistic | p-value | Cohen's $d$ |
| FCSRT: Total Free Immediate Recall | 350 | 0 | 19 (5.43) | -0.064 (1.046) | 0.048 (0.945) | -1.069 (1.354) | 4.752 | **< 0.001** | 1.125 |
| FCSRT: Total Immediate Recall | 350 | 0 | 30 (8.57) | -0.088 (1.132) | 0.045 (0.869) | -1.282 (2.134) | 3.647 | **< 0.001** | 1.251 |
| FCSRT: Total Free Delayed Recall | 350 | 0 | 27 (7.71) | -0.108 (1.09) | 0.027 (0.974) | -1.32 (1.319) | 5.868 | **< 0.001** | 1.33 |
| FCSRT: Total Delayed Recall | 350 | 0 | 28 (8) | -0.09 (1.205) | 0.068 (0.93) | -1.517 (2.142) | 4.335 | **< 0.001** | 1.431 |
| MBT: Total Immediate Paired Recall | 337 | 13 | 22 (6.53) | -0.049 (1.134) | 0.103 (0.956) | -1.397 (1.624) | 5.28 | **< 0.001** | 1.44 |
| MBT: Total Immediate Free Recall | 336 | 14 | 21 (6.25) | -0.048 (1.074) | 0.092 (0.967) | -1.328 (1.172) | 6.716 | **< 0.001** | 1.437 |
| MBT: Total Delayed Paired Recall | 337 | 13 | 25 (7.42) | -0.038 (1.178) | 0.12 (0.966) | -1.451 (1.819) | 4.958 | **< 0.001** | 1.455 |
| MBT: Total Delayed Free Recall | 334 | 16 | 23 (6.89) | -0.095 (1.113) | 0.035 (1.005) | -1.323 (1.334) | 5.589 | **< 0.001** | 1.305 |
| WMS: Total Immediate Recall | 335 | 15 | 22 (6.57) | -0.105 (1.029) | 0.014 (0.913) | -1.167 (1.358) | 4.949 | **< 0.001** | 1.223 |
| WMS: Total Delayed Recall | 336 | 14 | 18 (5.36) | -0.122 (1.074) | -0.009 (0.946) | -1.13 (1.543) | 4.147 | **< 0.001** | 1.098 |
| WMS: Total Recognition | 336 | 14 | 24 (7.14) | -0.098 (1.031) | -0.006 (0.949) | -0.911 (1.348) | 3.808 | **< 0.001** | 0.908 |

The table presents robust complex SRB change indices for the episodic memory battery. Development of these indices has been previously described (see López-Martos, D., et al., 2025). Robust longitudinal complex SRB indices reflect intra-individual cognitive change adjusted for demographics and referenced to the Aβ-negative sample (z-scores). For each variable, the table reports the number of observations (n), not available missing data (NA), the number and percentage (%) of individuals below the 5th percentile ( $< -1.645$ SRB), and the mean (SD) values for the full sample. Results are also stratified by group (stable, Amnesic obj-SCD), with mean (SD), t-statistic, p-value, and Cohen's $d$ for group comparisons. Bold text indicates significant results ( $p$ -values $< 0.05$ ). Aβ : β-amyloid; FCSRT: Free and Cued Selective Reminding Test; MBT: Memory Binding Test; SD: standard deviation; SRB: standardized regression-based; WMS: Wechsler Memory Scale.

### Neuropsychological Measurements

#### Tests and Questionnaires

##### Free and Cued Selective Reminding Test (FCSRT)

The Spanish FCSRT was used to assess episodic memory employing a controlled learning procedure.^35^ Participants learned 16 semantically unrelated words paired with cues and completed three recall trials, each preceded by a 20-second subtraction task. Trials included free recall followed by cued recall for unrecalled items, with selective reminding applied only in the first two trials. Delayed free and cued recall was assessed after 25–35 minutes. Variables of interest included total free immediate recall (0–48), total immediate recall (0–48), total free delayed recall (0–16), and total delayed recall (0–16).

##### Memory Binding Test (MBT)

The Spanish MBT was used to assess episodic memory employing an associative learning procedure.^36^ Participants learned two lists of 16 semantically paired words under controlled conditions, followed by recall of each list by semantic category and free recall of all 32 items. Delayed recall (free and facilitated) was tested after 30 minutes. Variables of interest included total immediate paired recall (0–32), total immediate free recall (0–32), total delayed paired recall (0–32), and total delayed free recall (0–32).

##### Wechsler Memory Scale–IV (WMS-IV) Logical Memory (LM) Subtest

The Spanish WMS-IV LM subtest was used to assess narrative episodic memory.^37^ Participants immediately recalled two orally presented stories (B and C), recalled them after 20–30 minutes, and then completed a recognition task. Variables of interest included immediate recall (0–50), delayed recall (0–50), and recognition (0–30).

##### Clinical Dementia Rating (CDR)

The Spanish CDR was used to screen global cognition and daily functioning across six domains, yielding a global score from 0 (no impairment) to 3 (severe dementia).^38^

##### Mini Mental State Examination (MMSE)

The Spanish MMSE was used to screen global cognition, assessing orientation, registration, attention, recall, language, and constructive praxis (0–30).^39^

##### Subjective Cognitive Decline Questionnaire (SCD-Q)

The Spanish SCD-Q was used to assess SCD, completed by both participant and study partner, independently.^40^ The questionnaire contains three initial yes/no items and 24 questions on everyday cognitive difficulties. Categorically defined SCD status was determined according to the first item (“Do you perceive memory or cognitive difficulties?” for participants; “Do you perceive he/she has cognitive or memory difficulties?” for study partners). Total scores quantified SCD (0–24).

#### Clinical Staging

##### Amnestic, objective subtle cognitive decline (obj-SCD)

Amnestic obj-SCD was defined a priori, using a longitudinal, multi-criteria psychometric approach selected to enhance diagnostic sensitivity and specificity. Participants were required to meet the following criteria: significant cognitive decline across multiple episodic memory measures (FCSRT, MBT, and WMS-IV LM variables previously defined), operationalized as AD biomarker-based complex SRB change < -1.645 SD per variable. Complex SRB change indices were referenced to the Aβ-negative group, adjusted for age, sex, and education using previously validated robust longitudinal neuropsychological references (ALFA+ cohort).^25^ A multivariate base rate threshold was applied to categorically determine the presence of amnestic obj-SCD: participants with ≥10 measures were required to show decline on ≥3 measures, and those with <10 measures were required to show decline on ≥2 measures.^41^ All participants had complete longitudinal data available for the 4 main measures of the FCSRT. Full details are provided in the Supplementary Material (Supplementary Table 3).

##### Amnestic, mild cognitive impairment (MCI)

Amnestic MCI was defined a priori, using a cross-sectional multi-criteria psychometric approach applied at both baseline and follow-up, selected to enhance diagnostic specificity and maintain consistency with standard clinical practice. Participants were required to meet the following criteria: CDR global score > 0; MMSE score < 27, and the conservative application of the neuropsychological criteria for aMCI,^42^ operationalized as performance < 1.65 SD below the conventional normative mean on at least 2 of 4 main measures from the FCSRT.

### Statistical Analysis

#### Characteristics of the participants

Baseline characteristics of the full sample of participants were summarized and characteristics of the two groups of interest (stable and amnestic obj-SCD) were compared using univariate ANOVAs for continuous variables and Chi-square tests for categorical variables. Nominal p-values < 0.05 were considered statistically significant.

#### Robust Normative Longitudinal Neuropsychological Change

Neuropsychological variables used for clinical staging were summarized descriptively and compared between groups (stable and amnestic obj-SCD) using t-tests, with effect sizes quantified by Cohen’s d. All statistical analyses were conducted in R (version 4.2.1) within RStudio (version 2022.07.1). Nominal p-values < 0.05 were considered statistically significant.

#### Fluid Biomarkers

Associations of amnestic obj-SCD with plasma and CSF biomarkers were evaluated using linear mixed-effects (LME) regression models (lme4; R 4.2.1, RStudio 2022.07.1). Log10-transformed biomarkers were modeled as outcomes in separate models, with group (0: Stable, 1: Amnestic obj-SCD) as the main predictor, adjusting for age, sex, *APOE-ε4*, and time. Models included two time points, a group × time interaction, and a participant-level random intercept. Time visit was coded categorically, with age modeled as time-varying to account for exact follow-up intervals. Results are reported as standardized β coefficients with 95% confidence intervals, *p*-values, marginal and conditional R², and AIC. Nominal *p*-values < 0.05 were considered significant. Full details are provided in Supplementary Material (Supplementary Table 4).

#### Neuroimaging Biomarkers

Associations of amnestic obj-SCD with PET and GMv biomarkers were assessed using voxel-wise fixed-effects regression models (SPM12; MATLAB 2022b). Neuroimaging data were modeled as outcomes in separate models, with group (0: Stable, 1: Amnestic obj-SCD) as the main predictor, adjusting for age, sex, *APOE-ε4*, TIV (GMv only), and follow-up interval in years (longitudinal models only). Models applied an explicit GM mask, further excluding the cerebellum. Results are reported at cluster and peak levels, including cluster size (k), *p*-values and *T*-values, peak MNI coordinates (x, y, z), and anatomical labels. Nominal *p*-values < 0.005 with cluster extent > 100 voxels were considered significant. Full details are provided in Supplementary Material (Supplementary Table 5).

#### Exploratory Analysis

Exploratory analyses, restricted to the subset of participants with amnestic obj-SCD, evaluated associations of log10-transformed fluid biomarkers with observed longitudinal changes in GMv using fixed-effects linear regression models (RStudio). Longitudinal GMv changes were extracted at the cluster level from ROIs defined by previous group-level contrasts (MarsBaR 0.45; MATLAB 2022b). Mean GMv change values normalized for TIV, were modeled as outcomes, adjusting for age, sex, and follow-up interval in years. Results are reported in line with previous models. Nominal *p*-values < 0.05 were considered significant. Full details are provided in Supplementary Material (Supplementary Table 6).

#### Data Availability

The data that support the findings of this study are available from the corresponding authors, D.L.-M., O.G.-R. or G.S.-B., on reasonable requests.

## Results

### Characteristics of the Participants

A total of 350 CU participants were included in the present study, with a mean longitudinal time interval of 3.30 years (standard deviation = 0.52). According to the clinical staging criteria, 315 individuals (90%) were classified as stable, and 35 (10%) as exhibiting amnestic obj-SCD, with no participant fulfilling criteria for amnestic MCI. Baseline characteristics of the participants are presented in **Table 1**. Significant baseline differences were found between stable and amnestic obj-SCD groups. The group exhibiting amnestic obj-SCD was slightly older (*p*-value < 0.001), demonstrated a greater prevalence of Aβ positivity (*p*-value = 0.002), higher plasma p-tau217 concentration (*p*-value = 0.009), higher CSF p-tau181/Aβ42 ratio (*p*-value < 0.001), higher Aβ PET load (*p*-value < 0.001), lower performance on MMSE (*p*-value = 0.048) and greater SCD-Q scores, considering both self-reported (*p*-value = 0.004) and study-partner reports (*p*-value = 0.003), independently. No significant baseline differences were found for sex, education, *APOE-ε4* status, and categorically defined SCD status, considering both self-reported and study-partner reports, independently. Within the subset of amnestic obj-SCD, categorically defined self-reported SCD was present in 15 out of 35 individuals (43%) at baseline, increasing to 20 out of 35 individuals (57%) at follow-up.

### Robust Normative Longitudinal Neuropsychological Change

Results of AD biomarker-based robust complex SRB change indices used for clinical staging are presented in **Table 2**. In the full sample, the prevalence of subtle decline per variable (SRB change ≤ 5th percentile) ranged from approximately 5% to 9% across FCSRT, MBT, and WMS measures. The highest prevalence rates were observed for FCSRT Total Immediate Recall (8.57%), FCSRT Total Delayed Recall (8.0%), FCSRT Total Free Delayed Recall (7.71%), MBT Total Delayed Paired Recall (7.42%), and WMS Total Recognition (7.14%). Overall, SRB change scores, expressed as demographically adjusted z-scores referenced to the Aβ-negative sample, ranged from approximately -0.12 to - 0.04 at the full-sample level, indicating minimal average longitudinal cognitive change across the cohort. Within the stable group, SRB change scores were generally positive and centered near zero, reflecting relative cognitive stability over the study period, with most measures showing minimal improvement. In contrast, SRB change scores in the amnestic obj-SCD group were consistently negative across all measures, generally below -1 SRB, with mean values ranging from approximately -0.91 to -1.52, reflecting longitudinal decline. Participants classified as amnestic obj-SCD exhibited significantly lower SRB change scores than the stable group across all episodic memory measures examined (all *p*-values < 0.001). The most pronounced negative mean values were observed for FCSRT Total Delayed Recall (−1.52), MBT Total Delayed Paired Recall (−1.45), and MBT Total Immediate Paired Recall (−1.40). The largest group differences were observed for MBT Total Delayed Paired Recall (Cohen’s d = 1.455), MBT Total Immediate Paired Recall (Cohen’s d = 1.440), and MBT Total Immediate Free Recall (Cohen’s d = 1.437), followed by FCSRT Total Delayed Recall (Cohen’s d = 1.431) and FCSRT Total Free Delayed Recall (Cohen’s d = 1.330).

### Associations of Amnestic obj-SCD with Fluid Biomarkers

Associations of amnestic obj-SCD with fluid biomarkers are presented in **Figure 1**. Considering core AD biomarkers, amnestic obj-SCD was associated with higher plasma p-tau217 concentration (β_STD_[95%CI]= 0.409 [0.123, 0.686]), higher CSF p-tau181/Aβ42 ratio (β_STD_[95%CI]= 0.637 [0.322, 0.953]), and greater longitudinal increases of CSF p-tau181/Aβ42 ratio over time (β_STD_[95%CI]= 0.219 [0.053, 0.384]), compared to the stable group. Considering biomarkers of non-specific processes involved in AD pathophysiology, amnestic obj-SCD was associated with higher plasma NfL concentration (β_STD_[95%CI]= 0.400 [0.076, 0.723]), higher CSF NfL concentration (β_STD_[95%CI]= 0.348 [0.058, 0.638]), higher plasma GFAP concentration (β_STD_[95%CI]= 0.564 [0.262, 0.865]), and higher CSF GFAP concentration (β_STD_[95%CI]= 0.324 [0.0.027, 0.621]), compared to the stable group.

**Figure 1.**
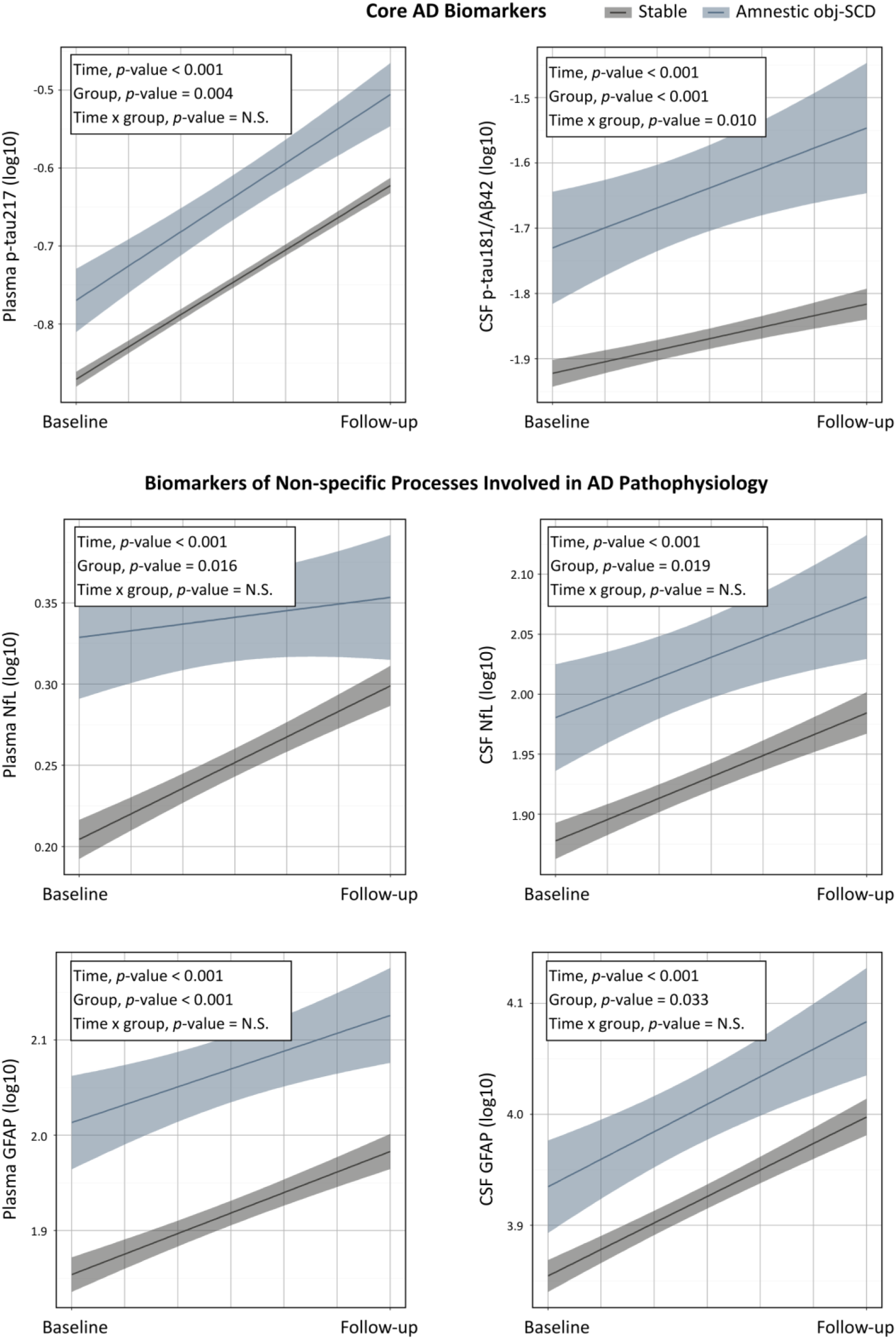
Associations of amnestic obj-SCD with Fluid Biomarkers. Results from LME regression models show the main effects of group, time, and group × time interaction on longitudinal plasma and CSF biomarker outcomes, including core AD markers and biomarkers of non-specific processes involved in AD pathophysiology. Shaded areas indicate 95% confidence intervals of the predicted regression lines. Aβ: β-amyloid; AD: Alzheimer’s disease; CSF: cerebrospinal fluid; GFAP: glial fibrillary acidic protein; NfL: neurofilament light chain; p-tau: phosphorylated tau.

### Associations of Amnestic obj-SCD with Neuroimaging Biomarkers

Associations of amnestic obj-SCD with neuroimaging biomarkers are presented in **Figures 2** and **3**. Considering core AD biomarkers, amnestic obj-SCD was associated with higher global Aβ PET load at baseline, predominantly in frontoparietal regions, relative to the stable group. Longitudinally, amnestic obj-SCD showed further increases in Aβ PET load, following a similar but more localized pattern of accumulation, most prominently in the precuneus. Amnestic obj-SCD was also associated with higher tau PET load, particularly in bilateral regions of the medial temporal lobe (MTL), compared to the stable group. Considering biomarkers of non-specific processes involved in AD pathophysiology, amnestic obj-SCD was associated with baseline reduced GMv, primarily in the middle cingulate cortex, with additional localized involvement of middle temporal, precentral, and postcentral regions. Longitudinally, amnestic obj-SCD was associated with GMv bilateral reductions in the hippocampus, alongside GMv increases in neocortical regions, primarily involving right-lateralized parietal and occipital regions, with additional, localized involvement of frontal and insular areas.

**Figure 2.**
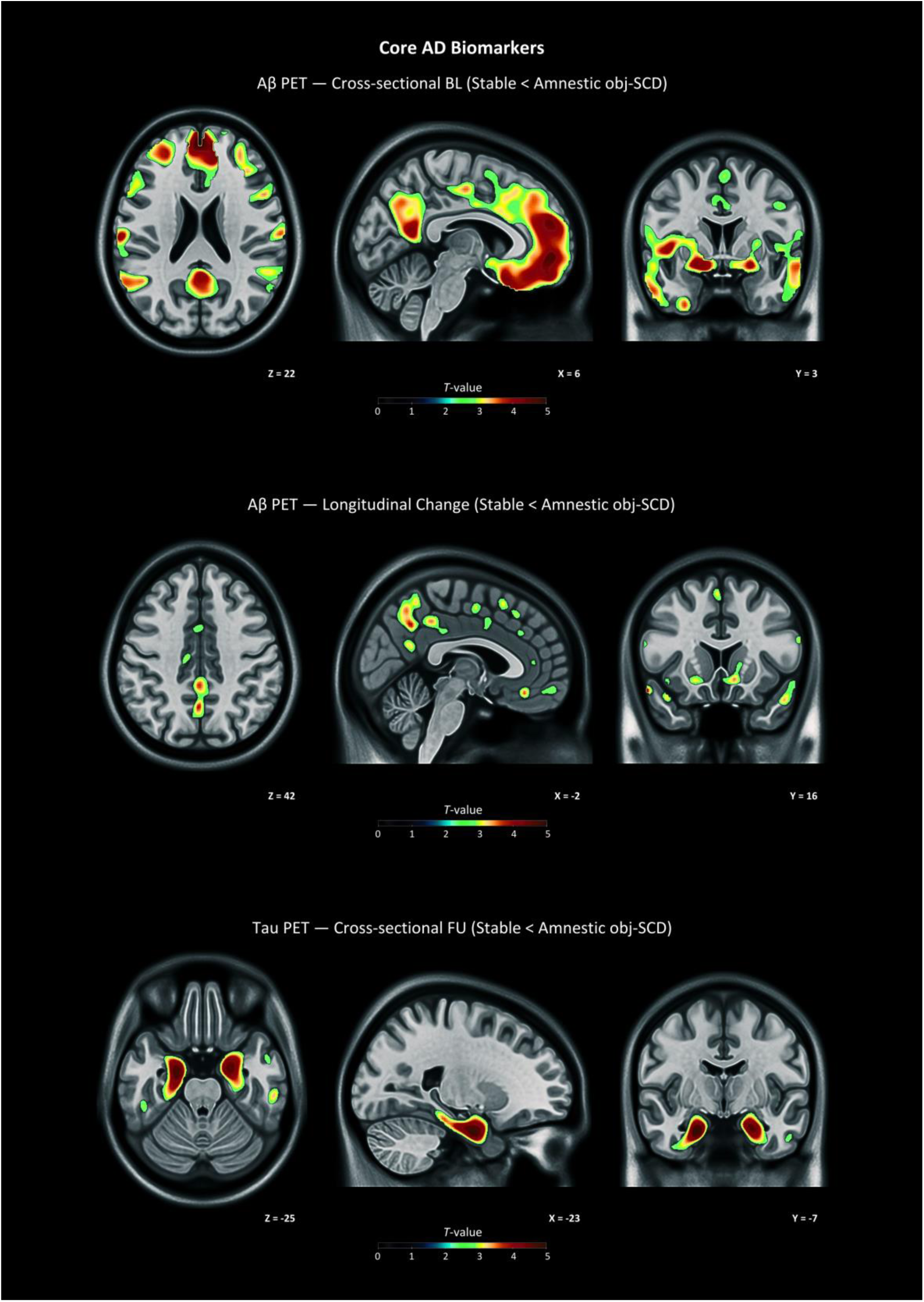
Associations of Amnestic obj-SCD with Neuroimaging Biomarkers: Core AD Biomarkers. Results from voxel-wise regression models show the association of amnestic obj-SCD with core AD biomarkers. The figure displays brain sections following anatomical convention, where the left side of the image corresponds to the left hemisphere of the brain. Color bars indicate *T*-values, with results shown at nominal *p*-values < 0.005 accounting for a cluster-size threshold of *k* > 100 voxels. Aβ: β-amyloid; AD: Alzheimer’s disease; BL: baseline; FU: follow-up; PET: positron emission tomography.

**Figure 3.**
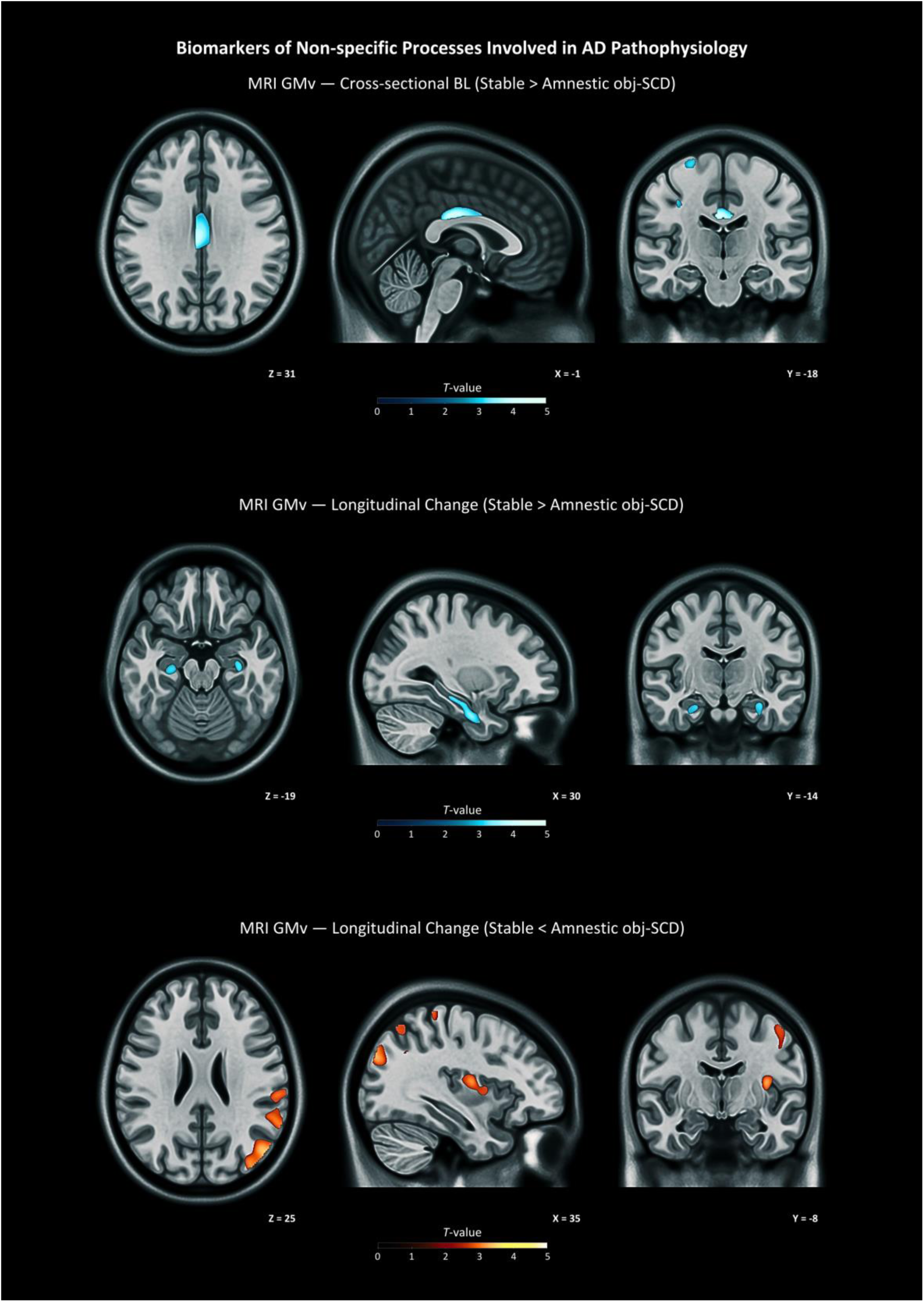
Associations of Amnestic obj-SCD with Neuroimaging Biomarkers: Non-specific Processes Involved in AD Pathophysiology. Results from voxel-wise regression models show the association of amnestic obj-SCD with biomarkers of non-specific processes involved in AD pathophysiology. The figure displays brain sections following anatomical convention, where the left side of the image corresponds to the left hemisphere of the brain. Color bars indicate *T*-values, with results shown at nominal *p*-values < 0.005 accounting for a cluster-size threshold of *k* > 100 voxels. AD: Alzheimer’s disease; GMv: gray matter volume; MRI: magnetic resonance imaging.

### Exploratory Analysis

Exploratory analysis was conducted restricted to the sample of participants with amnestic obj-SCD, associations of fluid biomarkers with longitudinal GMv change in ROIs are presented in **Figure 4**. ROIs were defined based on post-hoc, previous group-level contrasts, including ROIs showing longitudinal GMv reductions (bilateral hippocampus) and ROIs showing longitudinal GMv increases (neocortical regions). Within amnestic obj-SCD, higher baseline CSF p-tau181/Aβ42 ratio was associated with steeper longitudinal GMv reductions in bilateral hippocampus ROIs showing longitudinal GMv reductions (β_STD_[95%CI]= -0.369 [-0.647, -0.090]). In contrast, lower baseline plasma GFAP concentration (β_STD_[95%CI]= -0.401 [-0.781, -0.022]) and greater longitudinal increase in plasma GFAP concentration (β_STD_[95%CI]= 0.391 [-0.018, 0.764]) were both associated with steeper longitudinal GMv increments in neocortical ROIs showing longitudinal GMv increases.

**Figure 4.**
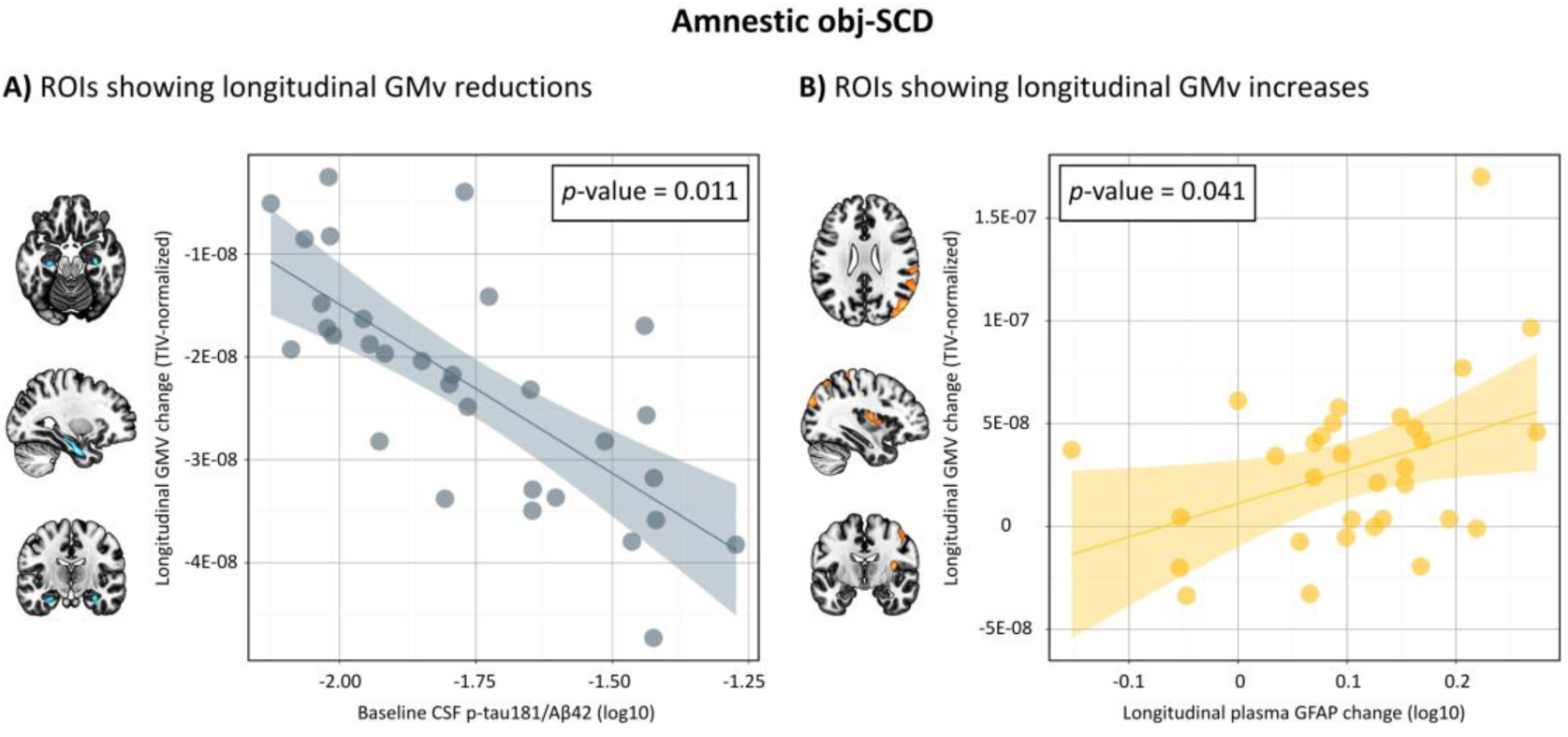
Associations of Fluid Biomarkers with Longitudinal GMv Changes in Amnestic obj-SCD. Results from linear regression models show the associations between fluid biomarkers and longitudinal GMv changes in amnestic obj-SCD, with shaded areas representing 95% confidence intervals of the predicted regression lines. Models were adjusted for age, sex, and longitudinal time interval measured in years (in models using longitudinal biomarker data). Panel A shows the negative association between baseline CSF p-tau181/Aβ42 and GMv change in ROIs showing longitudinal GMV reductions. Panel B shows the positive association between longitudinal change in plasma GFAP and GMv change in ROIs showing longitudinal GMV increases. Mean longitudinal GMv changes by ROIs were normalized for TIV. Aβ: β-amyloid; CSF: cerebrospinal fluid; GFAP: glial fibrillary acidic protein; GMv: gray matter volume; p-tau: phosphorylated tau; ROIs: regions of interest; TIV: total intracranial volume.

## Discussion

This study provided a comprehensive multimodal biomarker characterization of standardized longitudinal neuropsychological criteria for defining amnestic obj-SCD in aging and preclinical AD. The presence of amnestic obj-SCD was defined through robust AD biomarker-based longitudinal neuropsychological references, applying a multivariate base rate threshold of significant cognitive decline. According to the defined criteria, 35 individuals (10% of the present sample) exhibited amnestic obj-SCD over a 3-year longitudinal period. Multimodal biomarker characterization showed consistent significant associations of amnestic obj-SCD with core AD biomarkers, including diagnostic core 1 AD biomarkers (higher plasma p-tau217 concentration, higher CSF p-tau181/Aβ42 ratio, and higher global Aβ PET burden) and prognostic core 2 AD biomarkers (higher MTL tau PET burden). Additionally, amnestic obj-SCD was significantly associated with biomarkers of non-specific processes involved in AD pathophysiology, including neurodegeneration (higher plasma/CSF NfL concentrations, reduced GMv in the middle cingulate cortex, and longitudinal bilateral GMv reductions in the hippocampus) as well as inflammation (higher plasma/CSF GFAP concentrations and longitudinal GMv increases in neocortical regions). These findings suggest that amnestic obj-SCD reflects early AD-related neuropathological changes, alongside downstream mechanisms involved in AD pathophysiology, aligning with the integrated clinical-biological staging proposed by 2024 Alzheimer’s Association revised criteria.^4^ This study highlights the need to strengthen current frameworks through the development of objective, standardized criteria for clinical staging, supporting early detection and risk stratification in aging and preclinical AD.

Amnestic obj-SCD was defined with clinically grounded methods, selected to balance sensitivity and specificity. Neuropsychological assessment incorporated comprehensive, standardized, and challenging episodic memory tests, encompassing the main variables from the FCSRT, MBT, and WMS-IV LM. Robust neuropsychological references based on AD biomarkers, which measured deviation relative to the group with nonpathological biomarker levels, enhanced sensitivity for detecting subtle cognitive decline in preclinical AD.^25,43^ Neuropsychological assessment depends on the normative references applied, with growing evidence indicating that robust normative frameworks provide higher diagnostic standards than conventional references.^18,19^ The use of demographically adjusted complex SRB longitudinal change indices, reflecting deviation from normative intra-individual cognitive variability over time, allowed to apply a cut-off consistent with clinical practice to define statistically significant longitudinal decline at the variable level (*i.e.*, SRB change ≤ 5^th^ percentile). Neuropsychological methods tracking change relative to an individual’s baseline are essential, as cross-sectional assessments may be insufficient for detecting significant cognitive decline, particularly in the absence of clinical impairment. Within this framework, SRB models have been proposed as a sensitive approach for defining obj-SCD,^44^ and have been previously validated in both MCI and dementia stages.^45^ Finally, to distinguish AD-related transitional decline from normal aging, more advanced methods than those typically used for MCI diagnosis are required. Because diagnostic accuracy and stability are influenced by the number of impaired measures considered and specific cut-offs used,^46^ a multivariate base rate threshold of significant decline was employed to enhance specificity in amnestic obj-SCD determination, controlling false-positive classifications.^27^

Amnestic obj-SCD was associated with core AD biomarkers across fluid and neuroimaging modalities. Amnestic obj-SCD was significantly associated with fluid biomarkers, including higher plasma p-tau217 concentration and CSF p-tau181/Aβ42 ratio. Beyond the significant main group effect, both biomarker levels showed significant longitudinal increases across the full sample, with plasma p-tau217 exhibiting the greatest rise. However, only CSF p-tau181/Aβ42 demonstrated a significant group × time interaction, indicating greater longitudinal increase in amnestic obj-SCD. While the pronounced elevation in plasma p-tau217 indicated widespread early AD-related changes across the full sample, in consistency with its established sensitivity to underlying Aβ pathology in CU individuals as demonstrated in large population-based cohorts,^47–49^ the longitudinal increase in the CSF p-tau181/Aβ42 ratio showed a closer correspondence with the memory decline defining amnestic obj-SCD. This pattern is consistent with evidence indicating that plasma p-tau217 is well suited for scalable detection of early AD pathology, whereas CSF and PET provide greater specificity, relevant for integrated clinical-biological staging.^49^ These results align with previous studies linking core AD biomarkers to cognitive decline in CU population.^50,51^ Neuroimaging analyses further showed significant associations of amnestic obj-SCD with Aβ and tau PET. Baseline Aβ PET load was significantly higher in amnestic obj-SCD, with greater global burden and prominent frontoparietal deposition. Longitudinally, amnestic obj-SCD showed significant increases in Aβ PET load, most notably in the precuneus. Tau PET, acquired at follow-up, was also significantly higher in amnestic obj-SCD, with greater burden in bilateral MTL regions. These results were consistent with AD biomarker staging,^4^ implicating regions affected across early-intermediate Aβ progression^52^ and early MTL-restricted tau accumulation characteristic of clinical stage 2, prior to widespread neocortical involvement and overt clinical impairment.^53,54^ Together, these findings align with the established predictive value of multimodal core AD biomarkers for cognitive decline in CU populations and support their role in refining early clinical–biological staging,^50,55^ including characterization of clinical Stage 2, as reflected by amnestic obj-SCD.

Amnestic obj-SCD was also associated with biomarkers of non-specific processes involved in AD pathophysiology across fluid and neuroimaging modalities, including neurodegeneration and inflammation. Amnestic obj-SCD was significantly associated with fluid biomarkers, including higher neurodegeneration measured by plasma and CSF NfL concentrations, as well as higher inflammation measured by plasma and CSF GFAP concentrations. These results align with previous research linking neurodegenerative and inflammatory mechanisms with cognitive decline in CU population.^56,57^ Beyond the significant group effect, all these biomarker levels showed longitudinal increases across the full sample, with plasma GFAP exhibiting the greatest rise; however, there were no significant group × time interactions. This pattern suggests that neurodegenerative and inflammatory processes are active early in aging, but their longitudinal changes are not specific to amnestic obj-SCD. In contrast, longitudinal increases in CSF p-tau181/Aβ42 ratios and Aβ PET burden were directly associated with amnestic obj-SCD, highlighting a closer link between changes in these AD-specific biomarkers and clinical Stage 2. Neuroimaging analyses showed key differences in GMv. At baseline, amnestic obj-SCD was associated with reduced GMv, primarily in the middle cingulate cortex, with additional involvement of middle temporal, precentral, and postcentral regions. Longitudinally, amnestic obj-SCD showed bilateral GMv reductions in the hippocampus alongside GMv increases in neocortical regions. These findings align with the early vulnerability of the hippocampus in AD, supporting the objective characterization of a subclinical amnestic syndrome.^58–61^ Cingulate GMv reduction in posterior regions has been reported in preclinical AD,^62^ suggesting regional vulnerability in mid-posterior regions may emerge earlier than previously recognized, consistent with reports of cingulate atrophy in MCI.^63^ Finally, the observed longitudinal GMv increases in neocortex, although not well established in current AD frameworks, align with non-linear structural dynamics described across the preclinical stage.^64–67^

Exploratory analyses examined potential mechanisms underlying longitudinal GMv changes in amnestic obj-SCD. Results revealed a direct pattern of associations: the CSF p-tau181/Aβ42 ratio was associated with longitudinal GMv reductions in the bilateral hippocampus, whereas plasma GFAP concentration was associated with longitudinal GMv neocortical increases. These findings suggest core AD pathology contributes to the neurodegenerative mechanisms underlying the subclinical amnestic syndrome characterized with obj-SCD, consistent with evidence linking episodic memory decline to Aβ-related hippocampal atrophy,^68^ with longitudinal GMv increases in neocortical regions associated with astrocytic reactivity reflecting expected transient morphological changes in brain structure, potentially arising as an early inflammatory response to AD pathology.^69^ Nevertheless, further research is needed to clarify the mechanisms linking specific pathological processes to structural brain dynamics in aging and preclinical AD, particularly in relation to the emergence of objective, subclinical manifestations.^70,71^

Several aspects require further development to translate these findings into clinical practice, including validation across independent cohorts. As expected, the frequency of amnestic obj-SCD was low (10%), given the stringent criterion of longitudinal decline < 5^th^ percentile on multiple measures, aimed to characterize the lower end of neuropsychological distributions.^25^ Prognostic value of the proposed criteria should be determined with longer follow-up. Importantly, the operational definition of MCI, which delineates the boundary of obj-SCD, directly influences prevalence estimates and prognostic interpretation. Prevalence of obj-SCD across other domains (*e.g.*, dysexecutive, linguistic, mixed variants) should be addressed in future studies, as cognitive phenotyping distinguishing MCI subtypes have shown to influence the diagnostic performance of AD biomarkers.^72^ Clinically, obj-SCD may support earlier risk stratification but requires further validation before routine implementation, future work should explore shorter-term assessments within the 1-3 years window to enable wider research and clinical use. Diagnostic criteria and clinical staging should address meta-cognitive assessment to ensure conceptual alignment with the natural course of AD.^73^ In this study, individuals exhibiting amnestic obj-SCD demonstrated substantial heterogeneity in cognitive awareness (> 40% amnestic obj-SCD did not report SCD at follow-up), suggesting the emergence of subtle amnestic anosognosia.^74^ This finding underscores a key limitation of SCD for defining Stage 2, a critical yet often neglected clinical aspect. Previous studies suggest that self-reported SCD contributes to MCI misdiagnosis,^75^ while reduced cognitive awareness or anosognosia is associated with clinical progression across the AD *continuum*.^7,10^ The present study defined amnestic obj-SCD without requiring positivity for a core 1 AD biomarker, with only 60% of cases Aβ-positive at baseline. Although current diagnostic guidelines emphasize biologically confirmed AD, the 2024 revised criteria acknowledge substantial individual variability,^4^ recognizing that not all individuals follow a canonical temporal sequence of biological and clinical events,^76^ highlighting the need for further research on vulnerability and resilience mechanisms, as well as comorbid pathologies in AD.

In conclusion, this study evaluated objective, standardized criteria for defining amnestic obj-SCD using robust longitudinal neuropsychological references with multivariate base rate thresholds of significant cognitive decline, demonstrating significant associations with core AD biomarkers and biomarkers of non-specific processes involved in AD pathophysiology. These findings underscore the need to develop standardized clinical staging criteria, supporting early detection and risk stratification in aging and preclinical AD.

## Data Availability

The data that support the findings of this study are available from the corresponding authors, D.L.-M., O.G.-R., or G.S.-B., on reasonable request.

## Acknowledgments

This publication is part of the ALFA study. The authors would like to express their most sincere gratitude to the ALFA project participants and relatives without whom this research would not have been possible. The authors also thank Roche Diagnostics International Ltd for providing the kits used to measure CSF biomarkers, the laboratory technicians at the Clinical Neurochemistry Laboratory in Mölndal, Sweden, for performing the analyses, Eli Lilly and Company for providing the measurements of the in-house assay for plasma p-tau217, GE Healthcare for supplying the [¹⁸F]flutemetamol doses, and F. Hoffmann-La Roche Ltd for sponsoring the tau PET study.

COBAS and ELECSYS are trademarks of Roche. All other product names and trademarks are the property of their respective owners. The NeuroToolKit is a panel of exploratory prototype assays designed to robustly evaluate biomarkers associated with key pathologic events characteristic of AD and other neurological disorders, used for research purposes only and not approved for clinical use (Roche Diagnostics International Ltd, Rotkreuz, Switzerland). The Elecsys β-Amyloid(1–42) CSF and Elecsys Phospho-Tau (181P) CSF assays are approved for clinical use.

## Appendix

### Collaborators

Collaborators of the ALFA study are: Federica Anastasi, Annabella Beteta, Marta del Campo, Lidia Canals, Alba Cañas, Irene Cumplido-Marinero, Carme Deulofeu, Ruth Dominguez, Maria Emilio, Karine Fauria, Ana Fernández-Angulo, Sherezade Fuentes, Marina García, Patricia Genius, Armand González-Escolar, Laura Hernández, Felipe Hernández-Viadel, Gema Huesa, Jordi Huguet, Laura Iglesias, Esther Jiménez, Ferran Lugo, Paula Marne, Tania Menchón, José Luis Molinuevo, Paula Ortiz-Romero, Wiesje Pelkmans, Albina Polo, Sandra Pradas, Blanca Rodríguez-Fernández, Iman Sadeghi, Lluís Solsona, Anna Soteras, Laura Stankeviciute, Núria Tort-Colet, Marc Vilanova, Natalia Vilor-Tejedor.

