## Supplementary Material for "Multimodal biomarker characterization of amnestic objective subtle cognitive decline"

***Corresponding authors:**

#### Supplementary Material

#### Methods

##### Study Participants

This study was performed in the Alzheimer's and Families+ (ALFA+) cohort, a longitudinal extension of the *ALzheimer's and FAmilies* (ALFA) parent cohort at the Barcelonaβeta Brain Research Center, Barcelona, Spain (Molinuevo et al., 2016). The ALFA parent cohort included 2,743 CU individuals with a high proportion of AD patients' offspring (86% regardless parental age of onset) and apolipoprotein E (*APOE*) *ε4* carriers (35%). The nested ALFA+ study screened 451 participants and included 419 participants selected by their specific AD risk profile, considering AD parental history and *APOE-ε4* (Molinuevo et al., 2016). A detailed phenotyping of the participants, aside from clinical, cognitive, behavioral, and lifestyle characterization, involved blood and CSF sample collection, as well as MRI and PET acquisitions.

The ALFA+ inclusion criteria were: (1) individuals who had previously participated in the ALFA study; (2) age between 45 and 75 years at the inclusion in the ALFA parent cohort; and (3) long-term commitment to the study: agreement to undergo all tests and study procedures (MRI, PET, and lumbar puncture [LP]). ALFA+ exclusion criteria were: (1) cognitive impairment (Clinical Dementia Rating [CDR] > 0) or low cognitive performance (Mini-Mental State Examination [MMSE] < 27 or semantic fluency < 12); (2) any systemic illness or unstable medical condition that could lead to difficulty complying with the protocol; (3) any contraindication to any test or procedure; and (4) a family history of monogenic AD.

##### Neuropsychological Assessment

Administration Procedure

Participants were invited for longitudinal neuropsychological assessments. Episodic memory was assessed using the Free and Cued Selective Reminding Test (FCSRT), the Memory Binding Test (MBT), and the Wechsler Memory Scale–IV (WMS-IV) Logical Memory (LM) Subtest. In total, 11 episodic memory variables were included in the definition of amnestic objective subtle cognitive decline (obj-SCD): 4 variables from the FCSRT: total free immediate recall (0–48), total immediate recall (0–48), total free delayed recall (0–16), and total delayed recall (0–16); 4 variables from the MBT: total immediate paired recall (0–32), total immediate free recall (0–32), total delayed paired recall (0–32), and total delayed free recall (0–32); and 3 variables from the WMS-IV LM Subtest: immediate recall (0–50), delayed recall (0–50), and recognition (0–30). Global cognition was screened with Clinical Dementia Rating (CDR) and Mini Mental State Examination (MMSE). SCD was assessed with the Subjective Cognitive Decline Questionnaire (SCD-Q). Neuropsychological assessments were conducted over two sessions, at both baseline and follow-up visits, with sessions separated by approximately three months. For each visit, the FCSRT, WMS-IV LM subtest, CDR, MMSE, and SCD-Q were administered in the first session, while MBT was administered in the second session. Administration procedure was consistent across baseline and follow-up visits.

### Results

**Supplementary Table 1.** Fluid biomarker data

| **Measurements** | **Full Sample (N= 350)** | **Stable (N= 315)** | **Amnestic obj-SCD (N= 35)** |
|---|---|---|---|
| *Core AD biomarkers* | | | |
| Plasma p-tau217 | | | |
| Baseline | 349 | 314 | 35 |
| Follow-up | 338 | 303 | 35 |
| CSF p-tau181/Aβ42 | | | |
| Baseline | 350 | 315 | 35 |
| Follow-up | 264 | 238 | 26 |
| *Biomarkers of non-specific processes involved in AD pathophysiology* | | | |
| Plasma NfL | | | |
| Baseline | 348 | 313 | 35 |
| Follow-up | 333 | 299 | 34 |
| CSF NfL | | | |
| Baseline | 350 | 315 | 35 |
| Follow-up | 264 | 238 | 26 |
| Plasma GFAP | | | |
| Baseline | 348 | 313 | 35 |
| Follow-up | 333 | 299 | 34 |
| CSF GFAP | | | |
| Baseline | 350 | 315 | 35 |
| Follow-up | 264 | 238 | 26 |

The table presents the number of available observations for each fluid biomarker and time point (rows), with participant counts shown for the full sample and stratified by group (columns). Biomarkers include core AD markers (plasma p-tau217, CSF p-tau181/Aβ42) and non-specific processes involved in AD pathophysiology (plasma and CSF NfL, plasma and CSF GFAP). Baseline refers to measurements at study entry, and follow-up corresponds to assessments at the second visit (3-year follow-up). Aβ: β-amyloid; AD: Alzheimer's disease; CSF: cerebrospinal fluid; GFAP: glial fibrillary acidic protein; NfL: neurofilament light chain; p-tau: phosphorylated tau.

**Supplementary Table 2.** Neuroimaging biomarker data

| *Neuroimaging Biomarkers* | | | |
|---|---|---|---|
| **Measurements** | **Full Sample (N= 350)** | **Stable (N= 315)** | **Amnestic obj-SCD (N= 35)** |
| *Core AD biomarkers* | | | |
| Aβ PET | | | |
| Baseline | 293 | 265 | 28 |
| Follow-up | 189 | 174 | 15 |
| Tau PET | | | |
| Follow-up | 87 | 76 | 11 |
| *Biomarkers of non-specific processes involved in AD pathophysiology* | | | |
| MRI GMv | | | |
| Baseline | 327 | 295 | 32 |
| Follow-up | 300 | 269 | 31 |

The table presents the number of available observations for each neuroimaging biomarker and time point (rows), with participant counts shown for the full sample and stratified by group (columns). Biomarkers include core AD markers (Aβ PET, tau PET) and markers of non-specific processes involved in AD pathophysiology (MRI GMv). Baseline refers to imaging assessments at study entry, and follow-up corresponds to scans obtained at the second visit (3-year follow-up). Aβ: β-amyloid; AD: Alzheimer's disease; GMv: gray matter volume; MRI: magnetic resonance imaging; PET: positron emission tomography.

**Supplementary Table 3.** Multivariate base rate thresholds

| Multivariate base rate threshold (rules) | Stable | Amnestic obj-SCD | Total |
|---|---|---|---|
| Available measures ≥ 10 (Decline ≥ 3) | 302 | 33 | 335 (95.71%) |
| Available measures range from 4 to 9 (Decline ≥ 2) | 13 | 2 | 15 (4.29%) |
| Total | 335 (90%) | 35 (10%) | 350 (100%) |

The table presents multivariate base rate thresholds for cognitive decline. Rows indicate the rules applied to define the thresholds based on the number of available measures and the minimum number of significant longitudinal cognitive decline per variable required, and columns show participant classification by group (stable, amnestic obj-SCD). Values represent the number of observations and corresponding percentages (%).

**Supplementary Table 4.** Associations of Amnestic obj-SCD with Fluid Biomarkers

| Model outcome | n | Predictors | $\beta_{STD}$ (95% CI) | *p*-value | $R^2$ | AIC |
|---|---|---|---|---|---|---|
| *Core AD biomarkers* | | | | | | |
| Plasma p-tau217 | 349/338 | | | | 0.400/0.676 | -575.01 |
| | | Group | 0.409 (0.132, 0.686) | **0.004** | | |
| | | Time | 1.128 (1.024, 1.231) | **< 0.001** | | |
| | | Group x Time | 0.078 (-0.204, 0.361) | 0.586 | | |
| CSF p-tau181/Aβ42 | 350/264 | | | | 0.235/0.917 | -593.193 |
| | | Group | 0.637 (0.322, 0.953) | **< 0.001** | | |
| | | Time | 0.304 (0.223, 0.386) | **< 0.001** | | |
| | | Group x Time | 0.219 (0.053, 0.384) | **0.010** | | |
| *Biomarkers of non-specific processes involved in AD pathophysiology* | | | | | | |
| Plasma NfL | 348/333 | | | | 0.180/0.512 | -407.404 |
| | | Group | 0.400 (0.076, 0.723) | **0.016** | | |
| | | Time | 0.244 (0.118, 0.370) | **< 0.001** | | |
| | | Group x Time | -0.333 (-0.683, 0.018) | 0.063 | | |
| CSF NfL | 350/264 | | | | 0.343/0.887 | -988.165 |
| | | Group | 0.348 (0.058, 0.638) | **0.019** | | |
| | | Time | 0.367 (0.284, 0.450) | **< 0.001** | | |
| | | Group x Time | 0.029 (-0.161, 0.220) | 0.762 | | |
| Plasma GFAP | 348/333 | | | | 0.303/0.880 | -832.058 |
| | | Group | 0.564 (0.262, 0.865) | **< 0.001** | | |
| | | Time | 0.420 (0.338, 0.502) | **< 0.001** | | |
| | | Group x Time | -0.075 (-0.251, 0.100) | 0.400 | | |
| CSF GFAP | 350/264 | | | | 0.318/0.950 | -1228.204 |
| | | Group | 0.324 (0.027, 0.621) | **0.033** | | |
| | | Time | 0.744 (0.672, 0.815) | **< 0.001** | | |
| | | Group x Time | 0.068 (-0.060, 0.196) | 0.299 | | |

Results from mixed-effects regression models indicating model outcome, number of observations (n) at baseline/follow-up, standardized (STD) β coefficients, 95% Confidence Intervals (CI), nominal *p*-values, marginal/conditional $R^2$, and Akaike Information Criteria (AIC) measurements. The mixed-effects regression models used two time-points and included a random intercept for each participant. The structure of the independent models was the following: Biomarker ~ Age + Sex [0,1] + *APOE*-ε4 [0,1] + Group [0: Stable, 1: Amnestic obj-SCD] + Time [0: Baseline,1: Follow-up] + (Group [0,1] * Time [0,1]) + (1 | participant). Age was coded as a time-varying variable, accounting for the time interval between baseline and follow-up visits. Bold text indicates a significant association (*p*-values < 0.05). Aβ: β-amyloid; CSF: cerebrospinal fluid; GFAP: glial fibrillary acidic protein; NfL: neurofilament light chain; p-tau: phosphorylated tau.

**Supplementary Table 5.** Associations of Amnestic obj-SCD with Neuroimaging Biomarkers

| | | Cluster-level | | | Peak-level | | Peak-level MNI coordinates | | |
|---|---|---|---|---|---|---|---|---|---|
| **Model outcome, contrast** | **n** | **Anatomical location** | ***k*** | ***p*-value** | ***T*-value** | ***p*-value** | **x** | **y** | **z** |
| *Core AD biomarkers* | | | | | | | | | |
| Aβ PET — Cross-sectional BL | 293 | | | | | | | | |
| Stable > Amnestic obj-SCD | | | | | | | | | |
| | | - | - | - | - | - | - | - | - |
| Stable < Amnestic obj-SCD | | | | | | | | | |
| | | Middle Frontal Cortex Left | 33477 | **< 0.001** | 4.944 | **< 0.001** | -12 | 8 | -10 |
| | | Precuneus Right | 3218 | **< 0.001** | 4.308 | **< 0.001** | 2 | -56 | 16 |
| | | Middle Occipital Cortex Left | 288 | 0.138 | 4.006 | **< 0.001** | -46 | -72 | 4 |
| | | Middle Temporal Pole Left | 280 | 0.143 | 3.627 | **< 0.001** | -30 | 4 | -44 |
| | | Supramarginal Gyrus Right | 186 | 0.227 | 3.577 | **< 0.001** | 64 | -18 | 24 |
| | | Superior Temporal Gyrus Right | 361 | 0.100 | 3.440 | **< 0.001** | 56 | -48 | 18 |
| | | Calcarine Cortex Left | 155 | 0.269 | 3.430 | **< 0.001** | -6 | -96 | -6 |
| Aβ PET — Longitudinal change | 189 | | | | | | | | |
| Stable > Amnestic obj-SCD | | | | | | | | | |
| | | - | - | - | - | - | - | - | - |
| Stable < Amnestic obj-SCD | | | | | | | | | |
| | | Superior Temporal Cortex Left | 189 | 0.050 | 4.310 | **< 0.001** | -66 | -26 | 10 |
| | | Precuneus Right | 824 | **< 0.001** | 4.288 | **< 0.001** | 6 | -48 | 62 |
| | | Medial Orbitofrontal Cortex Right | 300 | 0.017 | 4.144 | **< 0.001** | 2 | 32 | -16 |
| | | Cuneus Right | 250 | 0.027 | 4.106 | **< 0.001** | 8 | -98 | 16 |
| | | Middle Frontal Cortex Left | 188 | 0.051 | 3.598 | **< 0.001** | -40 | 58 | -8 |
| | | Middle Temporal Cortex Left | 167 | 0.064 | 3.571 | **< 0.001** | -66 | -12 | -12 |
| | | Striatum Right | 130 | 0.097 | 3.565 | **< 0.001** | 12 | 18 | -8 |
| | | Middle Temporal Cortex Right | 314 | 0.015 | 3.509 | **< 0.001** | 70 | -24 | -12 |
| | | Putamen Left | 293 | 0.018 | 3.360 | **< 0.001** | -22 | 10 | -6 |
| | | Precuneus Left | 105 | 0.132 | 3.252 | **< 0.001** | -2 | -58 | 22 |
| Tau PET — Cross-sectional FU | 87 | | | | | | | | |
| Stable > Amnestic obj-SCD | | | | | | | | | |
| | | - | - | - | - | - | - | - | - |
| Stable < Amnestic obj-SCD | | | | | | | | | |
| | | Parahippocampal Gyrus Left | 1446 | 0.010 | 5.023 | **< 0.001** | -22 | -8 | -28 |
| | | Parahippocampal Gyrus Right | 1307 | 0.013 | 4.642 | **< 0.001** | 22 | -6 | -26 |
| *Biomarkers of non-specific processes involved in AD pathophysiology* | | | | | | | | | |
| MRI GMv — Cross-sectional BL | 327 | | | | | | | | |
| Stable > Amnestic obj-SCD | | | | | | | | | |
| | | Middle Cingulate Cortex Left | 741 | 0.054 | 4.718 | **< 0.001** | -4.5 | -22.5 | 27 |
| | | Middle Temporal Cortex Left | 352 | 0.17 | 3.673 | **< 0.001** | -43.5 | -52.5 | 18 |
| | | Precentral Gyrus Left | 360 | 0.165 | 3.172 | **< 0.001** | -31.5 | -7.5 | 55.5 |
| | | Postcentral Gyrus Left | 121 | 0.419 | 2.893 | **< 0.001** | -36 | -15 | 37.5 |
| Stable < Amnestic obj-SCD | | | | | | | | | |
| | | - | - | - | - | - | - | - | - |

| | | | | | | | | | |
|---|---|---|---|---|---|---|---|---|---|
| MRI GMv — Longitudinal change | 300 | | | | | | | | |
| Stable > Amnestic obj-SCD | | | | | | | | | |
| | | Hippocampus Right | 373 | 0.328 | 3.487 | **< 0.001** | 30 | -9 | -30 |
| | | Hippocampus Left | 332 | 0.356 | 3.437 | **< 0.001** | -25.5 | -18 | -22.5 |
| Stable < Amnestic obj-SCD | | | | | | | | | |
| | | Lingual Gyrus Right | 1889 | 0.037 | 3.917 | **< 0.001** | 9 | -57 | -3 |
| | | Angular Gyrus Right | 1294 | 0.078 | 3.562 | **< 0.001** | 48 | -70.5 | 28.5 |
| | | Insula Right | 233 | 0.442 | 3.215 | **< 0.001** | 36 | -9 | 12 |
| | | Superior Parietal Cortex Right | 1622 | 0.051 | 3.173 | **< 0.001** | 16.5 | -76.5 | 55.5 |
| | | Superior Parietal Cortex Left | 553 | 0.234 | 3.125 | **< 0.001** | -13.5 | -81 | 49.5 |
| | | Supramarginal Gyrus Right | 843 | 0.146 | 3.065 | **< 0.001** | 63 | -27 | 27 |
| | | Postcentral Gyrus Right | 588 | 0.221 | 2.973 | **< 0.001** | 28.5 | -39 | 64.5 |
| | | Precentral Gyrus Right | 143 | 0.555 | 2.809 | **< 0.001** | 45 | -9 | 54 |

The table presents associations of amnestic decline with neuroimaging biomarkers from whole-brain voxel-wise regression models. For each model outcome and contrast, the table shows the number of observations (n), cluster-level results (anatomical location, cluster size in voxels [*k*], cluster-level *p*-value), peak-level results (*T*-value, peak-level *p*-value), and peak-level coordinates in Montreal Neurological Institute (MNI) space. Anatomical locations were defined at the cluster-level, using 1 $mm^3$ resolution labeling from the Automated Anatomical Labelling Atlas 3. Voxel-wise models were structured as: Neuroimaging Outcome ~ Age + Sex [0,1] + APOE-ε4 [0,1] + Group [0: Stable, 1: Amnestic obj-SCD] + Total Intracranial Volume (for MRI GMv models) + Time Interval (for longitudinal models). Results are displayed at nominal *p*-values < 0.005 with a cluster-size threshold of *k* > 100 voxels. Bold text indicates significant associations (*p*-values < 0.005). AD: Alzheimer's disease; Aβ PET: β-amyloid positron emission tomography; Tau PET: tau positron emission tomography; BL: baseline; FU: follow-up; MRI GMv: magnetic resonance imaging gray matter volume.

**Supplementary Table 6.** Exploratory Analysis

| Model outcome | Predictors | | n | $\beta_{STD}$ (95% CI) | *p*-value | $R^2$ | AIC |
|---|---|---|---|---|---|---|---|
| *Longitudinal GMv in decrease ROIs* | | | | | | | |
| | Plasma p-tau217 | | | | | | |
| | | Baseline | 31 | -0.173 (-0.485, 0.140) | 0.267 | 0.493/0.415 | -1041.206 |
| | | Longitudinal change | 31 | 0.095 (-0.207, 0.398) | 0.523 | 0.476/0.396 | -1040.206 |
| | CSF p-tau181/Aβ42 | | | | | | |
| | | Baseline | 31 | -0.369 (-0.647, -0.090) | **0.011** | 0.586/0.522 | -1047.482 |
| | | Longitudinal change | 24 | -0.198 (-0.580, 0.183) | 0.289 | 0.499/0.393 | -801.130 |
| | Plasma NfL | | | | | | |
| | | Baseline | 31 | -0.274 (-0.607, 0.059) | 0.103 | 0.520/0.447 | -1042.940 |
| | | Longitudinal change | 31 | 0.226 (-0.072, 0.523) | 0.131 | 0.513/0.438 | -1042.479 |
| | CSF NfL | | | | | | |
| | | Baseline | 31 | -0.327 (-0.666, 0.012) | 0.058 | 0.538/0.466 | -1044.073 |
| | | Longitudinal change | 24 | -0.155 (-0.530, 0.220) | 0.397 | 0.488/0.380 | -800.604 |
| | Plasma GFAP | | | | | | |
| | | Baseline | 31 | -0.207 (-0.524, 0.111) | 0.192 | 0.502/0.425 | -1041.778 |
| | | Longitudinal change | 31 | 0.116 (-0.202, 0.435) | 0.460 | 0.479/0.399 | -1040.376 |
| | CSF GFAP | | | | | | |
| | | Baseline | 31 | -0.206 (-0.506, 0.093) | 0.168 | 0.506/0.430 | -1042.017 |
| | | Longitudinal change | 24 | 0.284 (-0.079, 0.647) | 0.118 | 0.533/0.435 | -802.840 |
| *Longitudinal GMv in increase ROIs* | | | | | | | |
| | Plasma p-tau217 | | | | | | |
| | | Baseline | 31 | -0.069 (-0.471, 0.333) | 0.726 | 0.163/0.034 | -922.462 |
| | | Longitudinal change | 31 | 0.033 (-0.350, 0.416) | 0.859 | 0.160/0.031 | -922.351 |
| | CSF p-tau181/Aβ42 | | | | | | |
| | | Baseline | 31 | 0.127 (-0.266, 0.521) | 0.511 | 0.173/0.046 | -922.836 |
| | | Longitudinal change | 24 | 0.158 (-0.328, 0.645) | 0.504 | 0.183/0.011 | -709.264 |
| | Plasma NfL | | | | | | |
| | | Baseline | 31 | -0.100 (-0.539, 0.340) | 0.645 | 0.166/0.038 | -922.570 |
| | | Longitudinal change | 31 | 0.073 (-0.317, 0.463) | 0.704 | 0.164/0.035 | -922.488 |
| | CSF NfL | | | | | | |
| | | Baseline | 31 | 0.112 (-0.344, 0.567) | 0.618 | 0.167/0.039 | -922.614 |
| | | Longitudinal change | 24 | -0.202 (-0.671, 0.267) | 0.378 | 0.198/0.029 | -709.692 |
| | Plasma GFAP | | | | | | |
| | | Baseline | 31 | -0.401 (-0.781, -0.022) | **0.039** | 0.289/0.179 | -927.492 |
| | | Longitudinal change | 31 | 0.391 (0.018, 0.764) | **0.041** | 0.287/0.177 | -927.414 |
| | CSF GFAP | | | | | | |
| | | Baseline | 31 | -0.206 (-0.588, 0.176) | 0.278 | 0.197/0.074 | -923.746 |
| | | Longitudinal change | 24 | 0.037 (-0.449, 0.523) | 0.876 | 0.164/-0.011 | -708.715 |

The table presents associations between fluid biomarkers and longitudinal GMv changes by decrease and increase ROIs within the subset of amnestic obj-SCD. Results from linear regression models show the number of observations (n), standardized β coefficients ($\beta_{STD}$) with 95% confidence intervals (CI), nominal *p*-values, $R^2$ (nominal/adjusted), and Akaike Information Criterion (AIC) values. Independent models were structured as: ROI ~ Age + Sex [0,1] + Biomarker + Time interval (years) for longitudinal models. Bold text indicates significant associations (*p*-values < 0.05). CSF: cerebrospinal fluid; GFAP: glial fibrillary acidic protein; GMv: gray matter volume; NfL: neurofilament light chain; p-tau: phosphorylated tau; ROIs: regions of interest.
